# Current approaches in livestock geotagging: Assessing available technologies and applications to public health research

**DOI:** 10.1101/2024.02.06.24302394

**Authors:** Julianne Meisner, Boku Bodha, J. Russell Stothard, Alexandra Juhasz, Peter Makaula, Janelisa Musaya, Isaac Ngere, John Gachohi, M. Kariuki Njenga, Dismas Oketch, Waiguru Muriuki, Eric Osoro

## Abstract

Livestock geotrackers are increasingly used for public health research, particularly within the field of One Health, to draw inference on pathogen exposure and human risk from livestock movement data. There are many dozens, if not hundreds, of devices available to researchers, including devices purpose built for livestock such as collars, ear tags, rumen boluses, or other formats; those intended for wildlife but suitable for livestock; and devices intended for other geotracking applications which can be retrofitted for livestock. To assist other researchers in navigating the wealth of available options, we present here our experiences with six devices—four intended for livestock, one intended for wildlife, and one intended for humans— each applied to cattle, camels, sheep, goats, and donkeys. We summarize the technical specifications and features of these devices, our deployment strategies, and our experiences in terms of battery life, durability, data quality and retrieval, and acceptability by livestock owners.

## INTRODUCTION

Satellite-based Global Positioning System (GPS) technology has been available to civilians since the 90s. Due to decreasing cost and increasing functionality, uptake of GPS technology is almost ubiquitous use today. Concurrent increase in computing power and adoption of so-called “Big Data” approaches to scientific research across a range of disciplines has led to widespread use of GPS technology to track livestock movements using wearable devices such as collars, ear tags, and rumen boluses. Ecologists use such methods to track spatiotemporal patterns of resource use and behavioral responses to environmental features, allowing resulting ecosystem impacts and species interactions to be modeled (1,2). In agricultural research, GPS devices are central to precision farming programs in industrialized systems, and are used to characterize the movements of transhumant and nomadic pastoralists in extensive systems (3).

In public health, livestock movements play a pivotal role in the epidemiology of various zoonotic diseases. For example, the role of livestock movement in the spread of Rift Valley fever (4) and *Taenia solium* (5) highlights the important connections between livestock mobility and the spread of infectious diseases. Epidemiologists have used livestock movement data to infer exposure to pathogens by coupling movement data with data on vector distributions (6) and human movements (7), and combining movement livestock and wildlife movement data to model wildlife-livestock contact networks (8). An understanding of the dynamic relationships between livestock movement and pathogen transmission is necessary for implementation of effective public health interventions and disease control strategies, and the integration of GPS tracking with Geographic Information Systems (GIS) has allowed for enhanced data analysis and informed decision-making.

Livestock geotagging has evolved markedly over the past decades. From rudimentary tagging systems to sophisticated GPS-based technologies, these advancements have transformed how livestock movements are monitored and analyzed (9). Researchers interested in using GPS devices now have a range of products to choose from, which have trade-offs in terms of cost, durability, weight, battery life, accuracy, data richness (variables collected), data storage, network coverage, and frequency of fixes, which itself has trade-offs with battery life. Users can purchase purpose-built products intended for wildlife or livestock tracking; retrofit products intended for recreation or asset tracking by attaching them to collars, with or without external battery packs; or custom build dataloggers using pre-assembled boards and open-access platforms such as Arduino (10).

Despite technological advancements, several challenges remain in effective livestock tracking. These include the cost of deploying GPS devices, their durability under different environmental conditions, and concerns related to data privacy and security. Additionally, the effectiveness of these technologies is often limited in specific contexts, such as dense forests, savannah, or remote regions, posing significant hurdles in data accuracy and reliability. Existing literature provides limited insights into the comparative performance of different GPS devices in diverse geographical settings.

Because the options for livestock GPS tracking are changing so rapidly, there is a need to periodically update the literature on available options and their trade-offs. We present here our collective experience using livestock GPS devices for public health, across three research projects conducted in pastoralist communities in Kenya, and subsistence livestock-keeping communities in Malawi from 2022 to 2023. The objective of this study is to evaluate and compare the performance and suitability of various GPS devices for livestock geotagging in these settings. By focusing on these specific geographical areas and a range of technological solutions, the study aims to provide actionable insights for optimizing livestock tracking in similar contexts. Across all three projects, a total of six commercially available devices were used: one device primarily manufactured for wildlife tracking, four devices manufactured for livestock security on ranches, and one device manufactured to monitor the safety of adults with dementia. We present specifications for each device; our protocols for their deployment; and our experience with each device type, including functionality, durability, and community acceptability.

## METHODS

### Study 1: human-animal contact networks in Marsabit, Kenya

#### Background and setting

Authors Julianne Meisner and Boku Bodha are the lead investigators on a study of human-animal contact networks in northern Kenya. Study communities include one agropastoralist Rendille community near Marsabit town, and two pastoralist communities in Marsabit county: one Borana, and one Gabra. These communities rear cattle, sheep, goats, donkeys and camels, separately in fora and home herds.

Data collection was conducted from December 2022 – July 2023. The onset of data collection coincided with the end of a three-year long drought in Marsabit during which many livestock were lost, resulting in marked reductions in herd size. At the time of data collection, mean herd sizes were 7.65 camels (range 0-60), 7.65 cattle (range 0-29), and 30.71 sheep and goats (range 0-147). When the drought broke in late March, heavy rains and flooding occurred in the county and dramatically altered livestock movements.

#### Sampling strategy

We used a network-based sampling strategy called ego-centric sampling, whereby initially-selected households (“egos”) are asked to name other households whose herds contact their own via shared water, grazing, husbandry, or breeding (“alters”). For this pilot study, we sampled one ego household and up to four alter households in each community, leading to a final sample of 15 households.

As our goal was to pilot both devices and methods for measuring human-animal contact, our deployment strategy oversampled ego herds so that we could use the generated data to simulate subsampling, and ultimately examine the number of animals per herd that should be tracked to minimize bias. We similarly oversampled the fora herd as the animals in this herd do not cluster as tightly as those in the home herd. Our strategy was as follows:

- Ego households: 24 devices divided evenly between camels, cattle, and sheep/goats. If the household keeps both a fora and home herd, 2/3rds of the devices should be placed in the fora herd, and the remaining 1/3^rd^ in the home herd.
- Alter households: 6 devices in total, divided evenly between camels, cattle, and sheep/goats, and the fora and home herds. If the household keeps a separate home and fora herd, and each herd includes members of each species group (camels, cattle, sheep/goats), then one device would be placed per species group in each herd.

Decisions regarding the duration of GPS tracking are guided by the total number of devices available (i.e., budget), battery life, and study logistics. There is a tradeoff between using a GPS device to track a single animal for a longer period of time, vs. several animals “sharing” that device by removing and re-deploying it. The choice between these two strategies should be guided by the research question and study logistics: study duration, labor required to remove and re-deploy devices, etc. Ultimately, for this study we opted to track livestock for four weeks, remove, and re-deploy on the next ego network. This fit with our goal of tracking a large number of animals in each ego herd (which necessitated re-use of devices), and the five months available for data collection for this pilot project.

#### GPS devices

This study piloted three types of livestock GPS devices, summarized in Table 1: FindMy, Digitanimal, and FarmRanger.

**Table 1:** GPS devices used in Study 1.

|  | *FindMy* | *Digitanimal* | *FarmRanger* |
| --- | --- | --- | --- |
| **Country of origin** | Norway | Spain | South Africa |
| **Networks** | Mobile phone, satellite | GSM, SigFox | GPRS |
| **App** | iOS; Android soon | iOS or Android | iOS or Android |
| **Subscription costs** | Per fix | First year included | Billed monthly |
| **Weight** | 200gm | 265gm | 226gm |
| **Size** | 75mm x 60mm x 55mm | 104mm x 76mm x 48mm | 120mm x 60mm x 45mm |
| **Collar material** | Flexible plastic | Woven canvas | Woven canvas |
| **Battery life** | 2000+ messages | 1-1.5 years for SigFox<br>6-7 months for GSM | 5-6 months with standard use, 3 months with hourly alerts |
| **Battery** | Sourced from manufacturer | Sourced from manufacturer | Rechargeable, cable included |
| **Number tested** | 18 | 20 (10 GSM, 10 SigFox) | 10 |

FindMy (https://findmy.no/en/agtech), based in Norway, produces e-bells for sheep, cattle, reindeer, camels, horses, and goats. Flexible plastic collars are sized to species, however the devices are interchangeable. Models are available which use satellite networks, mobile networks, or both. Service costs are available as subscription programs, or pay-as-you go message plans. Locations, battery status, inactivity alarms, distress alerts, and mortality events can be viewed in real time on a mobile app. Users can also use the app to customize preferences including message plan schedules and geofences. Batteries last over 2,000 fixes, but need to be purchased from the supplier.

Digitanimal (https://digitanimal.com/?lang=en), based in Spain, produces GPS devices for livestock. Woven canvas collars use counterweights to keep devices in the correct position on the left side of the neck. Large and small canvas collar sizes are available to fit small and large ruminants, however the devices and weights are interchangeable. Devices can be purchased for GSM (mobile) or SigFox (satellite) networks. Device costs include a one year subscription service for fixes every 30 minutes. Users can use a mobile phone app to customize a geofence and view temperature (current, weekly, and current pack), distance (current, weekly, and current pack), activity, and location history. Users can also customize alerts such as activity, loss or theft, temperature, exit from or entrance into a geofence, etc. Batteries last one to 1.5 years for SigFox and 6-7 months for GSM, but need to be purchased from the supplier.

FarmRanger (https://farmranger.co.za/), based in South Africa, manufactures GPS devices for cattle, sheep, goats, horses, and other animals for security purposes. Devices use mobile networks (GPRS), and subscription costs are billed monthly per device. Alerts are triggered if animals are stolen or attacked, or the device is removed or tampered with. Both the woven canvas collar and the setting of the GPS device is customized to animal species: if the device will be moved from cattle to sheep, the technical team must be alerted so the sensitivity can be adjusted. Sensitivity also changes between field and kraal/fora. Batteries last 5-6 months in standard functioning, where fixes occur every 30 seconds while in alarm mode, and the unit shuts off otherwise to conserve battery life. With hourly fixes, batteries are estimated to last 3 months. If sensitivity settings are incorrect, alarms will be triggered erroneously and battery life will be reduced. Batteries can be recharged using the provided USB Micro-B charger. No geofencing function is offered, only abnormal movements.

#### GPS fix rate

The fix rate we selected was ultimately influenced by study goals, available budget, and functionality of the devices we selected. In general, as fix rate frequency decreases (or errors increase), modeling and/or assumptions are required to infer contact events from GPS data. Generally speaking, for GPS data (c.f., other technologies such as proximity loggers), contact is defined by the occurrence of one or more fixes within a spatiotemporal envelope. For direct contact, this envelope is very small, corresponding to co-occurrence in the same space and time, and generally only allows for errors in positional accuracy and acquisition delays. For indirect contact, this envelope can be larger, corresponding to environmental persistence or dissemination (e.g., for pathogens with a free-living life stage).

Because livestock behavior is fairly predictable and easily observed compared with carnivores or wildlife, we expect relatively infrequent fix rates are sufficient to capture indirect contact. Livestock movements are characterized by diurnal rhythms—towards pasture and water in the morning and shelter in the evening—and influenced but not entirely determined by human keepers. We set fix rates to be every 30 minutes for the FarmRanger and Digitanimal devices, and five times per day for the FindMy devices: 01:00, 07:00, 10:00, 15:00, 19:00.

### Study 2: Role of camels and other livestock in the transmission of *Brucella* and Middle East respiratory syndrome coronavirus to humans; Marsabit and Kajiado counties, Kenya

#### Background and setting

The goal of the study is to use animal movement data to define the interactions between livestock herds that facilitate disease transmission, among pastoralist communities in Marsabit and Kajiado counties in Kenya. Author Eric Osoro is the lead investigator, collaborating with authors Kariuki Njenga, Isaac Ngere, John Gachohi, Dismas Oketchf, and Waiguru Muriuki.

Marsabit County, situated in Kenya’s arid northern region, is the home to many nomadic pastoralist communities, primarily the Gabra, Borana, and Rendille tribes. These communities heavily depend on their livestock (camels, cattle, sheep, and goats) for sustenance and economic stability. Conversely, Kajiado County, located in the semi-arid rangeland to the south of the country, is predominantly inhabited by the Maasai nomadic pastoralist community, who keep cattle, goats, and sheep.

Study 2 is a longitudinal cohort study conducted over 12-18 months, with data collection on human participants and their livestock every three months. Throughout the study’s duration, enrolled households receive visits at three-month intervals. Data collection began in December 2022 and is currently ongoing.

#### Sampling strategy

It is common practice in these communities for animals to be herded communally within specific geographical areas. In addition, distinct herding methods for camels, bovines, and sheep and goats result in varying movement patterns. Consequently, in consultation with community leaders, we took deliberate measures to select herds that were representative of movements for each species within their respective villages. We then attached a GPS collar to one animal within each identified herd, for all villages in our study area until each village had a GPS logger affixed to a representative member of each species group.

Once an animal received a collar, we kept it on that animal for the entire duration of the study. However, in cases where an animal was lost or sold, the collar could be transferred to another animal of the same species within the herd. Between May and July of 2023 we deployed a total of 81 GPS loggers across the two study sites: 20 Savannah GPS loggers applied to cattle and camels, and the remaining 61 CatLog2 GPS loggers distributed among all four livestock species under investigation.

#### GPS devices

This study tested two types of livestock GPS devices, summarized in Table 2: CatLog2 GPS logger, and Savannah GPS logger.

**Table 2:**
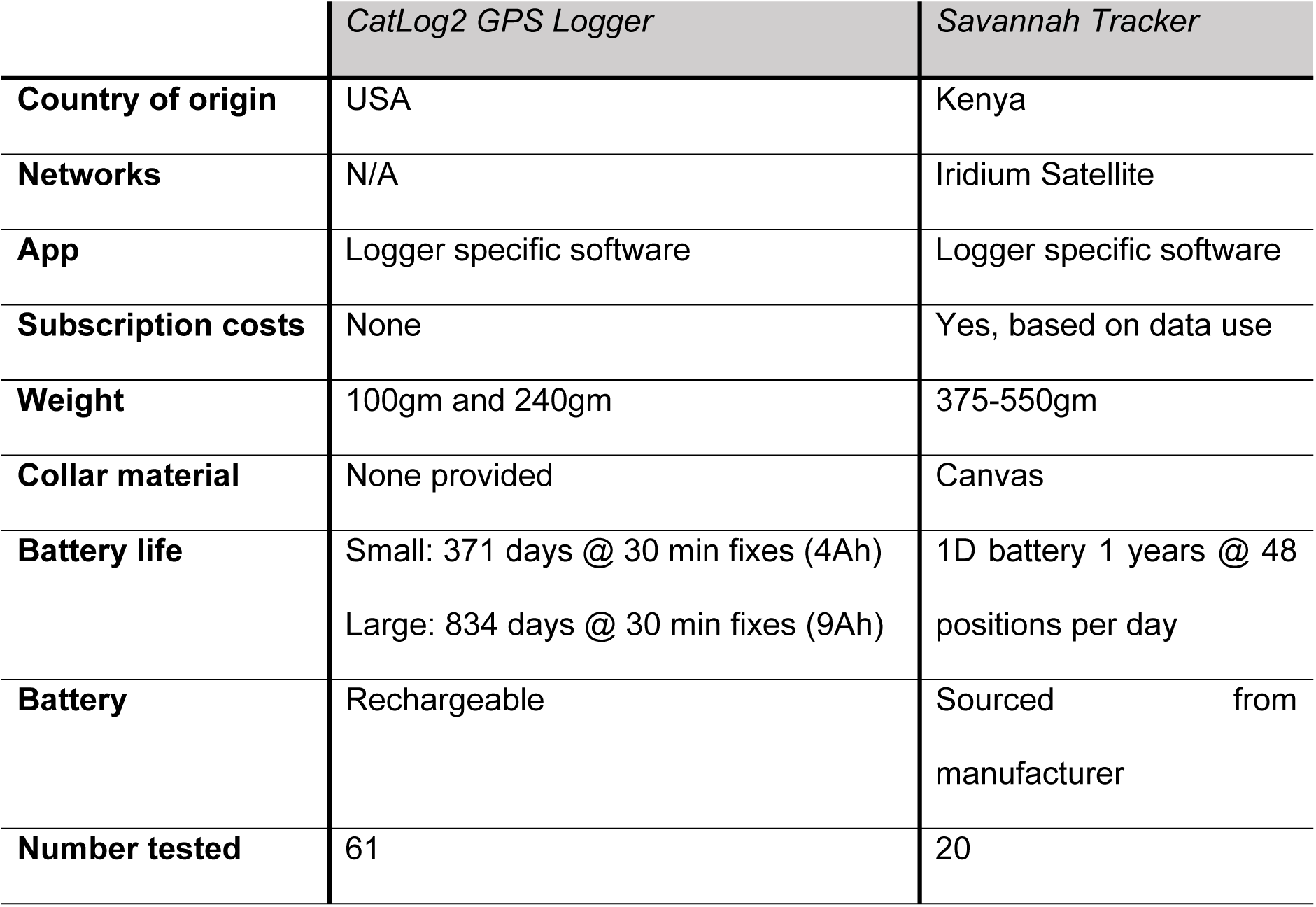
GPS devices used in Study 2.

|  | *CatLog2 GPS Logger* | *Savannah Tracker* |
| --- | --- | --- |
| **Country of origin** | USA | Kenya |
| **Networks** | N/A | Iridium Satellite |
| **App** | Logger specific software | Logger specific software |
| **Subscription costs** | None | Yes, based on data use |
| **Weight** | 100gm and 240gm | 375-550gm |
| **Collar material** | None provided | Canvas |
| **Battery life** | Small: 371 days @ 30 min fixes (4Ah)<br>Large: 834 days @ 30 min fixes (9Ah) | 1D battery 1 years @ 48 positions per day |
| **Battery** | Rechargeable | Sourced from manufacturer |
| **Number tested** | 61 | 20 |

The CatLog2 GPS logger, (http://www.mr-lee.com/catlog.htm), is housed within a tubular structure that combines a rechargeable battery and the GPS receiver. A collar or other means of affixing the device to an animal must be customized. The GPS logger does not have GSM or satellite connection capabilities for data transmission, necessitating manual retrieval of data by removing the device from the collar. During data retrieval, a charged battery can replace a depleted one, extending the duration of data collection. The data are downloaded using CatLog GPS Call Center v.4.1 software, and exported as a .csv file. Data fields includes date, time, position (longitude, latitude, and altitude), temperature, height, and speed (in meters per second). We deployed two types of devices which differed by battery size. For larger animals (cattle and camels) we used the 9Ah battery variant, while for smaller animals (sheep and goats) we deployed a lighter device with a 4Ah battery. We secured the devices to animals using a leather collar— tailored to animal size—with a pouch to hold the logger and facilitate easy removal for data download and subsequent reattachment.

Savannah Tracker GPS loggers (Savannah Tracking Ltd, Kilifi, Kenya) are designed for wildlife and livestock monitoring. Both the battery and GPS unit are affixed to a canvas collar, and the battery system is equipped with a non-rechargeable 1D battery. These GPS loggers use the Iridium satellite system for data communication, enabling precise tracking of movements even in remote and challenging terrains. Their design allows the loggers to endure harsh environmental conditions, including extreme temperatures and exposure to water, ensuring the dependable collection of field data. Battery life is generally long, but depends on the selected sampling interval. For example, when configured with 30-minute intervals, 24 hours of active GPS tracking per day, a 30-second time-to-position lock (GPS time out), and a 24-hour interval for data reporting, the estimated battery life is approximately 335 days for the 1D cell. A comprehensive data management software package is provided, facilitating various functions such as general queries, integration of auxiliary information, and remote data downloads in CSV, KMZ, and SHP formats. Prior to deployment, both the CatLog2 and Savanna GPS loggers required activation and configuration of the GPS fix rates. In addition, the Savanna GPS logger offered the capability of remote adjustment of its GPS fix rate.

#### GPS fix rate

Our selection of the GPS fix rate was influenced by device specifications, expert guidance, and the study’s duration. We aimed to deploy the GPS devices for 12 to 15 months to capture seasonal variations in livestock movements between the rainy and dry seasons. In seeking expert advice, we consulted with researchers experienced in the use of GPS loggers in similar research contexts. We opted for 30-minute intervals as this would preserve battery life, ensuring that the Savannah Tracker batteries could sustain the study period lasting 12 to 15 months.

As part of the quality assessment of the position data, we calculated the fix success rate by dividing the number of recorded fixes by the total number of scheduled fixes, for data collected during July and August 2023.

The Savannah Tracker devices have the GPS timeout and “retry” configuration settings, where it retries taking a position every 90 seconds until it is successful. This is important considering the movement of the animals through the shrubs and under the forest canopy, which blocks the device’s access to satellite view. Additionally, the CatLog GPS logger offers speed-triggered interval adjustment.

### Study 3: schistosomiasis exposure in Mangochi, Malawi

#### Background and setting

Authors Russell Stothard and Janelisa Musaya are the lead investigators for this study, collaborating with authors Alexandra Juhasz and Peter Makaula. The primary aim is to investigate hybridization in human schistosomes arising from close contact with animal schistosomes. Principally a closed prospective cohort study of humans with environmental surveillance of freshwater intermediate snails, this project additionally allocated significant research effort towards investigating the putative zoonotic spillover of schistosomiasis into local livestock. This brings together a multidisciplinary team inclusive of various stakeholders engaged in cattle farming and husbandry. Set within a One Health approach, a key focus is upon estimating cattle water contact points and immersion times along this shoreline (see www.lstmed.ac.uk/hugs) to quantify transmission of *Schistosoma haematobium* and *Schistosoma mattheei* along Lake Malawi, Mangochi District within a calendar year. Data were collected from April to December of 2022.

#### Sampling strategy

The primary goal of the GPS datalogging sub-study was to estimate water contact events. We targeted a small cattle herd of 8 animals in which a hybrid schistosome was confirmed. Over a three-month period the movement of each animal was estimated. The data were analyzed to obtain a cartographic representation of the comparative dispersion of animals, and water immersion times, as a broad estimation of transmission (Juhasz et al. *in preparation*).

This cattle herd was kept each night in a household kraal. The movements of these eight animals were recorded from May 2022 to July 2022. Given the small herd size and its composition of calves and adults, all animals were tracked to give an estimation of water contact immersion times for each animal.

#### GPS devices and fix rate

The GPS trackers we used were adapted from dementia care technologies (https://www.techsilver.co.uk/product/satellite-gps-tracker/). This is a satellite-based system that does not require a mobile signal to locate the animal quickly and easily. It provides regular updates on any detected movement of the herd and alerts a mobile phone with updates every 5 minutes. It is possible to subscribe for location fixes every 2 minutes but this increases subscription costs and reduces battery life. The GPS tracker is small (H 6.83 cm x W 5.13 cm x D 2.14 cm) and lightweight (90g with batteries). It uses four AAA batteries and if the battery power is low, a warning alert gives 2-3 days notice.

To fit the GPS unit to each animal, a nylon collar with plastic buckle and cross-stitched cradle for were bought from Amazon. The nylon fabric uses high-density webbing to increase durability and the quick release buckle was made of sturdy plastic, which is lighter than metal buckles and was more comfortable to wear. Before we put the GPS tracker in the cradle, we wrapped a moisture-wicking material around it and put it in a plastic bag. Each GPS collar and tracker was marked with white paint with the Roman numerals (I-VIII).

## ETHICAL APPROVALS

Ethical approval and Animal Care and Use committee approval was obtained from relevant authorities in the US, UK, Kenya, and Malawi. Informed consent was documented in writing and witnessed in all three studies. Special attention was given to the attachment and use of GPS devices on animals to minimize discomfort and avoid harm; devices were lightweight and non-invasive, ensuring little interference with the animals’ natural behavior.

## RESULTS

### Study 1: human-animal contact networks in Marsabit, Kenya

#### FindMy

We found these devices were easy to put on, and while a bit more difficult to remove, this reflected the security of the fastener. The plastic collar was pliable and lightweight, and appeared comfortable to the animals. One device broke when it was kicked by another animal, but the remaining 17 devices were intact and functioning at the end of the sampling period. A photo of these collars is presented in Figure 1.

**Figure 1:**
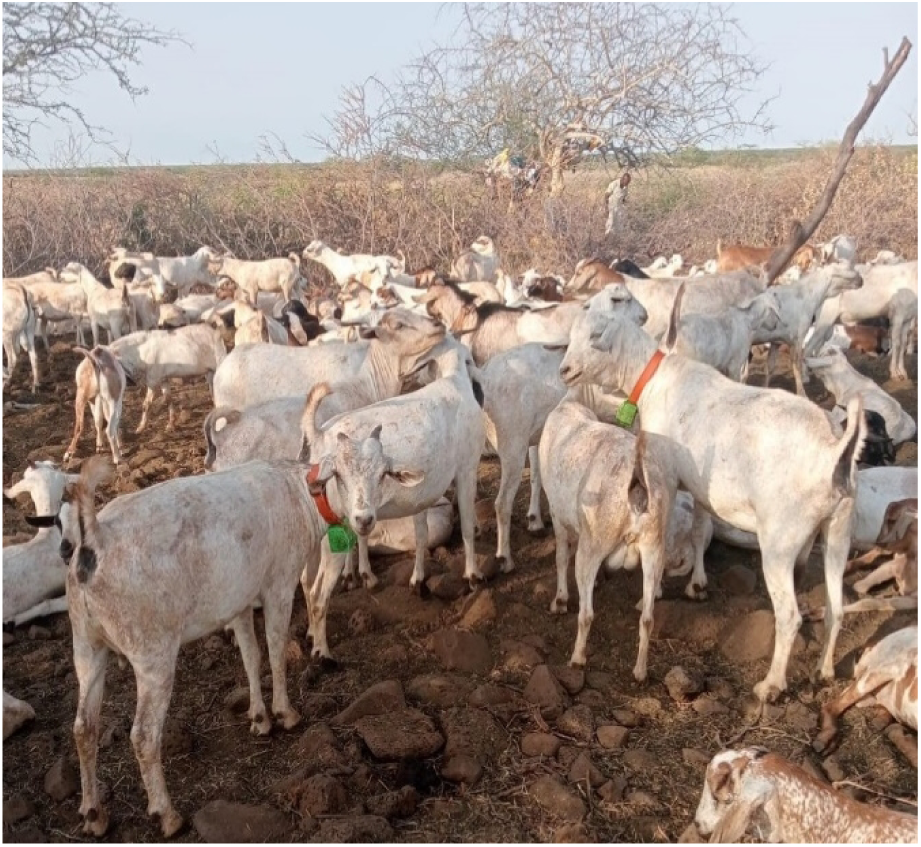
Goats in Marsabit wearing FindMy collars

Accuracy was estimated to be 19 meters, evaluated by calculating the positional error over 20 fixes compared to a handheld GPS device. Mean precision was 6.51 meters (range 0.5, 16.6) over the same 20 fixes. From the app, a total of 3,186 successful fixes were obtained, corresponding to a mean fix rate of 2.31 fixes/day, or a 41% fix rate success. From on-board storage, a total of 7,244 successful fixes were obtained, corresponding to a mean fix rate of 4.19 fixes/day, or an 83.7% fix rate. The missing fixes were all from the 01:00 fix time. We did not detect any implausible fixes, defined as fixes located outside of the subcounty. Observed battery life matched that expected.

#### Digitanimal

We found these devices were sturdy but complicated to attach to the animal. Both our study team and the community felt the device combined with the counterweight was too heavy for sheep and goats, particularly as the drought had weakened livestock in this region. Thus, we only applied these devices to cattle and camels. A photo of these collars is presented in Figure 2.

**Figure 2:**
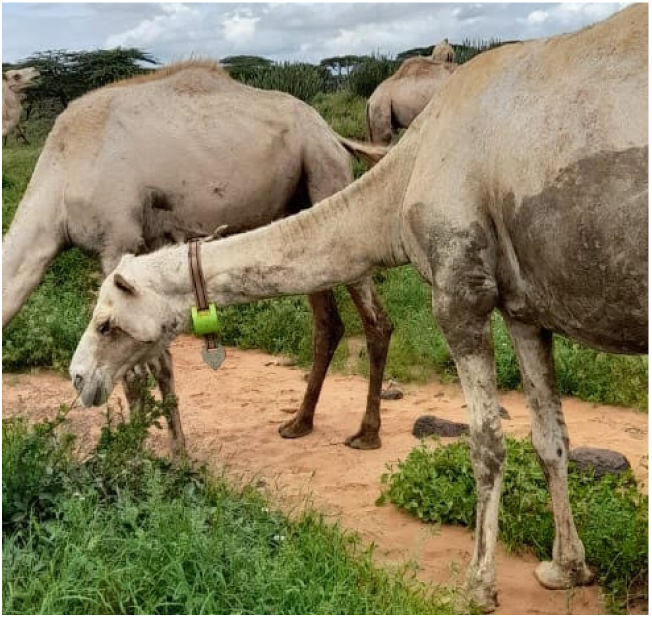
Camel in Marsabit wearing a Digitanimal collar

None of the SigFox devices worked in Marsabit. Of the 10 GSM devices, three stopped working between March and May of 2023. From the remaining seven GSM devices, accuracy was estimated to be 67 meters, estimated from 294 fixes. Mean precision was 94 meters (range 0, 943). A total of 10,794 successful fixes were obtained, corresponding to a mean fix rate of 31.1 fixes/day, or a 64% fix rate success for the devices that were still functioning. We did not detect any implausible fixes. Batteries lasted just under 6 months.

#### FarmRanger

We found that community members were enthusiastic about the design and appearance of these devices. For livestock keepers based in Africa wishing to collar their own animals, these devices are easy to ship within Africa, and can be recharged in the same way as a mobile phone. A photo of these collars is presented in Figure 3

**Figure 3:**
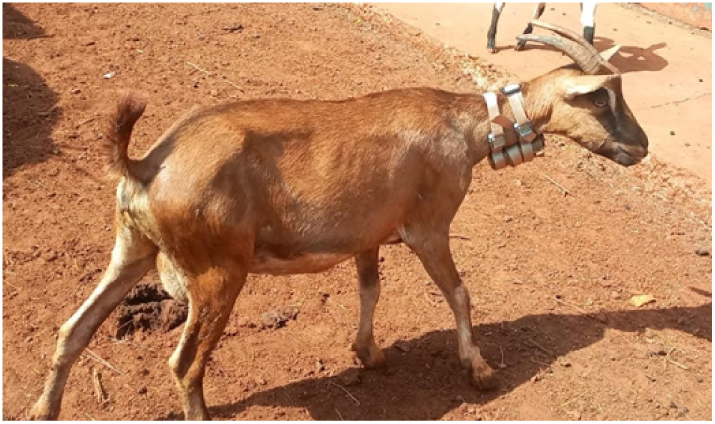
Goat in Marsabit wearing a FarmRanger collar

Accuracy was estimated to be 22 meters, from 18 fixes, and mean precision was 15.23 meters (range 0, 81). A total of 23,065 successful fixes were obtained, corresponding to a mean fix rate of 35.9 fixes/day, or a 74.7% fix rate success. We did not detect any implausible fixes. Batteries lasted just under 4 weeks, significantly shorter than expected for fixes every 30 minutes.

### Study 2: Brucellosis and MERS-CoV study

#### Savannah Tracker

Affixing the Savannah Tracker GPS logger was straightforward due to its adjustable neck collar. When restraining an animal, the collar was secured using flat washers and nuts, tailored to the animal’s neck girth. Care was taken to ensure it was neither too tight, causing discomfort, nor too loose, which could result in entanglement in stumps or tree branches during grazing. The collars exceeded the neck girth of the livestock, and any excess collar material was secured with plastic zip-ties to prevent skin irritation.

**Figure 4:**
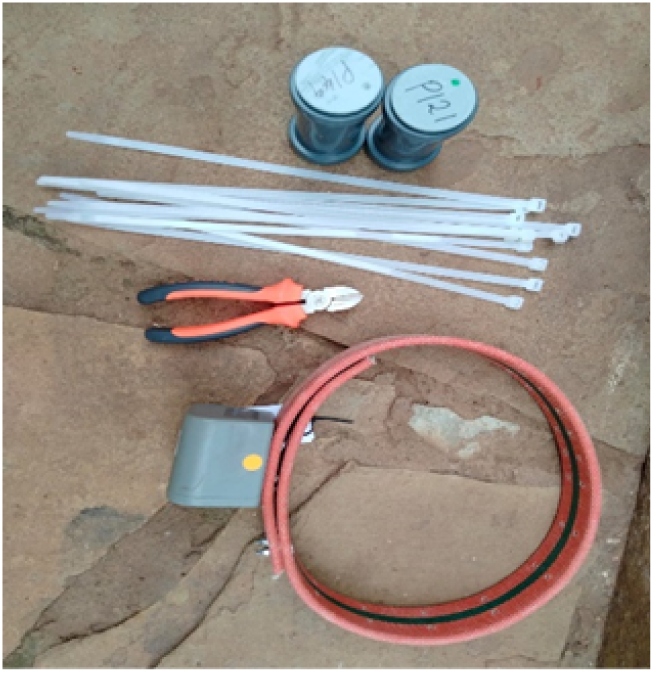
CatLog2 GPS logger (top) and Savannah GPS logger (bottom), with the items used for affixing the loggers.

Out of the 20 Savannah Tracker devices deployed on animals at the two study sites, two instances occurred where herders removed the devices. In one case, it was due to perceived discomfort to the animals, but the study team addressed this by resizing and redeploying the collar. In the second case, the household removed the device when they decided to de-stock the animal, subsequently contacting the study team for further action. Fix rate success was 99.7%.

#### CatLog2 GPS logger

For the CatLog2 GPS loggers, locally crafted collars were fashioned from animal leather, with sizes tailored for goats, sheep, cattle, and camels. These collars featured an integrated pouch designed to safeguard the devices against physical damage during the animals’ grazing activities. The attachment of CatLog2 GPS devices was facilitated by the use of zip-ties, which were secured around the pouch to affix the device firmly to the collar.

**Figure 5:**
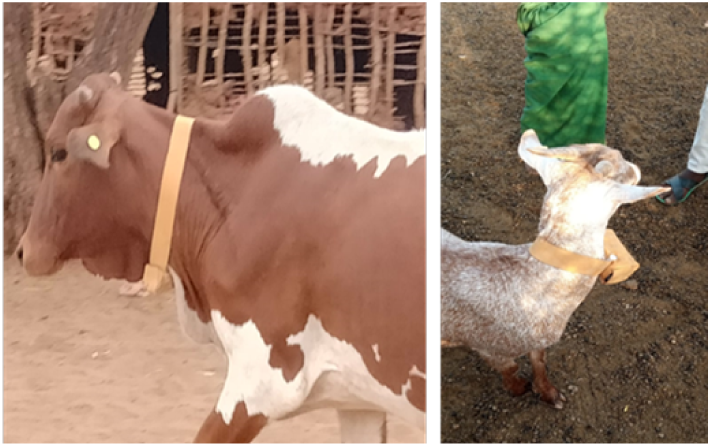
CatLog 2 GPS collar on a bovine (left) and goat (right)

Throughout the study, there were five instances in which CatLog2 devices became detached from animals, primarily camels and cattle. Two cases were due to wildlife attacks of livestock. The study team promptly reattached the devices to otheranimals within the same herds. The design of the GPS tracker and battery attachment left room for wire breakage during vigorous shaking, a situation that can arise when large-bodied animals engage in fights. In one instance, both the GPS tracker and batteries had to be replaced, while in another case, only the batteries required replacement. Fix rate success was 98.9%.

### Study 3: schistosomiasis exposure in Mangochi, Malawi

This satellite GPS tracker was ideal for tracking animals in rural areas, in close contact with humans. It caused minimal discomfort and did not change the animals’ behavior. Cattle in this setting are in regular contact with their owners, with their movements broadly controlled by a herder who is responsible for grazing and watering the animals under his supervision. Every second week, owners had to change the batteries, and without their help we would not have been able to do this study. In addition to the labor required to change batteries, our nylon straps started to fail as animals caught the material on trees and barbed wire fencing.

The manufacturer guaranteed their waterproofing, to a depth of 1m for up to 30 minutes. Nevertheless, the GPS tracking devices were not fully waterproof. If the animal stayed in the water for a long time, the moisture penetrated the device and caused false distances.

**Figure 6:**
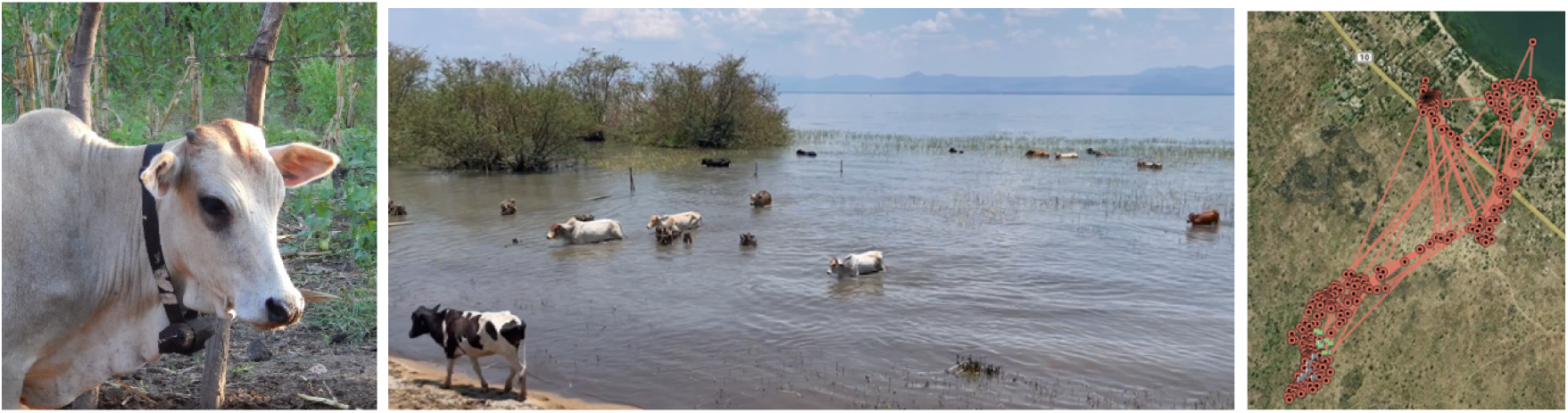
Cattle in Malawi wearing a satellite GPS collar (left) and cattle in water browsing on emergent vegetation often fully immersing the GPS device each day (middle) and plot of cattle movements over a 2-day period (right).

## DISCUSSION

Two studies (Study #1 and Study #2) were conducted among pastoralist livestock keepers in northern Kenya. This setting is characterized by spatiotemporal uncertainty in resource distribution, which pastoralist systems manage through seasonal movement.

In Study #1 we found that SigFox satellite coverage was not sufficient in Marsabit for the DigitAnimal SigFox devices to work. Reliability was also poor for the DigitAnimal GSM (mobile phone network) devices: several devices stopped working entirely, and the others had a low rate of successful fixes. This may reflect the setting itself, rather than the quality of the devices: we encourage other researchers to purchase 1-2 of the devices they plan to use to pilot them in their study setting. We found that acceptability was low due to the difficulty in attaching the collar, and the device’s weight, particularly for sheep and goats. Both the FarmRanger and FindMy devices were well-received. The FindMy devices had a much better battery life than FarmRanger, but FarmRanger devices are half as expensive, are easy to recharge, and can be easily shipped within Africa for farmers who wish to use them to track their own livestock. FarmRanger had a high fix rate success without need for collar retrieval to download data; FindMy had a low fix rate success for the app-based data, but a high fix rate success for the data stored on-board, which require collar retrieval to access. The app-based data were sufficient to use these to find the collars for retrieval and data download. As for DigitAnimal, this may reflect the setting rather than the device. Costs scale with the number of units purchased; we purchased between 10 (FarmRanger) and 20 (DigitAnimal) devices for $125-$260 per device, being the least expensive for FarmRanger, and the most expensive for FindMy. Precision and accuracy were sufficient for all three devices for our research questions, and all three devices were found to be extremely sturdy. Data collection has recently concluded, and analyses are currently being conducted.

In Study #2, we found the Savannah Tracker was extremely robust, accurate, and reliable, however the significant cost of these trackers (∼$1400) means they are most useful when the study objectives can be met with only a few animals tracked at one time. This can mean a small number of animals is tracked continuously, or a small number of collars are rotated between several (or many) animals. The CatLog2 tracker, by contrast, is far less costly ($100), but data must be manually retrieved, though battery replacement and data retrieval can be done at the same time. The Savannah Tracker was more sturdy than CatLog2, but CatLog2 still performed well in this regard. Both the Savannah Tracker and CatLog2 tracker had much higher fix rate success than the DigitAnimal, FindMy, or FarmRanger. Data collection is ongoing, and no analyses have been conducted yet.

Study #3 was conducted among subsistence farmers in Malawi, with cattle being moved to grazing and watering sites daily, and returning home each evening. This setting is more amenable to devices that require more frequent battery changes or manual data retrieval, and cellular network coverage is generally better. Of note, these devices were not manufactured for use on livestock and while they are fairly sturdy, they did not hold up as well as the other five devices discussed. Nonetheless, the observed geospatial tracks provide new insights into daily and seasonal cattle water contact patterns in Lake Malawi. These will be discussed in more detail in a subsequent publication; broadly, we found that during the dry season, animals spend up to 45 minutes immersed in shoreline water and frequently go into deep water to forage on emergent vegetation. We also found that the spatial patterns of livestock schistosomiasis transmission closely overlap with that of human urogenital schistosomiasis, indicating that livestock contaminate coastal waters and infect freshwater snails, which in turn infect humans who directly use lake water. Fully-assembled in the nylon collar and housing, these devices cost £162 each and have played crucial role in understanding the spatial epidemiology of bovine schistosomiasis and zoonotic spillover potentials

Livestock GPS tracking is increasing in popularity due to the growing availability of both devices, and analytic methods to analyze the data that result. While the iGot-U device has been extremely popular historically, it is increasingly less available, leaving researchers to navigate the many dozens (if not hundreds) of other options available. To assist other researchers interested in using these technologies we summarize our experiences with six devices here, linked to each study’s setting and objectives. With the exception of the Savannah Tracker, all of these devices cost at or under $250 per unit, making them accessible to research projects with a range of budgets. As the availability and functionality of livestock GPS technologies are likely to evolve rapidly, an important future direction is a platform or database where users can share their experiences with livestock GPS technologies for public health research. In selection of appropriate GPS devices for livestock tracking for public health research, we propose that researchers consider the cost of device, ease of application, retrieval of data, durability and battery life guided by the study setting.

Based on our findings, future research should focus on developing GPS devices with enhanced data transmission capabilities in low network areas and creating cost-effective solutions for widespread adoption. Collaborative efforts between technology developers, public health researchers, and local communities could lead to innovations that address the current limitations and widen the scope of GPS technology in livestock tracking for public health research.

## Data Availability

This manuscript presents our collective experiences using livestock GPS devices for public health research, including analysis of GPS functionality. These data are currently being analyzed to develop manuscripts related to the main hypotheses driving the respective studies, and will be released with those manuscripts.

